# Dry Needling in Parkinson’s Disease: A Randomized Clinical Trial with Markerless Kinematic Analysis

**DOI:** 10.64898/2026.03.10.26348054

**Authors:** Ariany Klein Tahara, Abel Gonçalves Chinaglia, Rafael Luiz Martins Monteiro, Lennin Abrão Sousa Santos, Paulo Roberto Pereira Santiago

## Abstract

Parkinson’s disease (PD) is associated with debilitating motor symptoms, including gait impairments and stooped posture. While markerless motion capture offers a scalable alternative to quantify these deficits, the acute biomechanical effects of interventions like Dry Needling (DN) on PD gait remain under-investigated. This randomized clinical trial investigated the acute effects of upper trapezius myofascial release via DN on gait and turning biomechanics in individuals with PD, utilizing a 3D markerless motion capture system. Thirty-eight participants with mild-to-moderate PD and bilateral upper trapezius trigger points were randomly assigned to a DN or Sham group. Gait was evaluated during a modified Timed Up and Go (TUG) test at baseline, immediately post-intervention, and at a one-week follow-up. Video data were processed using the open-source *vailá* toolbox, which applied a rigorous spatial segmentation algorithm to isolate postural transitions, steady-state gait, and turning phases. Thirty-seven participants completed the protocol (DN = 18; Sham = 19). The algorithm-driven markerless pipeline successfully extracted high-fidelity spatiotemporal parameters and automated gait event detection. However, the quantitative analysis revealed no significant Group × Time interactions for spatiotemporal parameters or turning kinematics, with both groups exhibiting similar trajectories across all assessment points. In conclusion, a single session of upper trapezius DN does not yield superior acute improvements in gait or turning biomechanics compared to a sham intervention in PD, suggesting that macro-level motor adaptations likely require cumulative therapeutic sessions. Nevertheless, the successful implementation of this markerless workflow provides an objective, cost-effective framework for precisely tracking phase-specific kinematic changes in routine clinical settings.

## INTRODUCTION

Parkinson’s disease (PD) is a progressive neurological disorder with the fastest-growing prevalence worldwide, characterized by a complex array of motor and non-motor symptoms. It affects movement primarily due to muscle rigidity and bradykinesia, leading to gait impairments and postural instability. [1, 2, 3] Slower walking speed, difficulty initiating gait, reduced stride rate, stride length variability, absence of arm swing, leg dragging, impaired turning, and a stooped posture are among the most common gait-related abnormalities observed in early PD, some of which may partially precede the formal diagnosis. [4]

Management and objective monitoring of these gait impairments remain a constant challenge in clinical settings. To address this, computational resources—particularly machine learning (ML) and deep learning (DL) approaches—have been increasingly applied not only to aid in early detection but, crucially, to track motor progression and treatment responses with high precision. [5]

Specifically, the integration of ML techniques with markerless pose estimation has advanced the capability to analyze human gait patterns. Machine learning models can process pixel-level information to predict spatiotemporal and kinematic gait parameters, providing a cost-effective and accessible alternative for motion analysis. This methodology enables reliable gait evaluation across diverse environments, overcoming the physical and financial limitations imposed by traditional marker-based motion capture systems. [6, 7]

To address the musculoskeletal consequences of the stooped posture and rigidity in PD, the dry needling technique (DNT) presents a promising therapeutic intervention. DNT utilizes fine filiform needles to deliver mechanical stimuli through the skin, targeting trigger points (TPs) within muscles and connective tissues. While there are currently no universally accepted criteria, such as biomarkers, electrodiagnostic testing, or imaging, for identifying or quantitatively describing TPs [8, 9, 10], evidence indicates that DNT produces immediate clinical benefits. These include pain reduction, increased range of motion (ROM), and potentially improved muscle activation through the treatment of active TPs. Given its clinical efficacy, accessibility, and favorable cost-effectiveness profile, DNT has been widely adopted in the management of various musculoskeletal disorders. [8, 11, 12, 13, 14] Notably, even a single DNT session has been shown to alleviate pain and improve ROM in conditions such as neck and shoulder pain. [15,16,17,18]

Therefore, the present study investigates the acute repercussions of upper trapezius myofascial release via DNT on gait kinematics in individuals with Parkinson’s disease. The primary objective is to assess the effectiveness of markerless ML approaches in detecting subtle, phase-specific gait pattern modifications induced by the intervention. Furthermore, this study aims to explore how objective markerless assessment can contribute to the clinical management and development of personalized therapeutic strategies for individuals with PD

## METHODOLOGY

### Patients, interventions and instruments

Patients aged over 50 years, with a clinical diagnosis of idiopathic Parkinson’s disease (PD) according to the Hughes criteria [19,20] classified as having mild to moderate PD (modified Hoehn and Yahr scale scores between 1.5 and 3.0), using levodopa, able to walk without assistive devices and maintain an upright posture, capable of following verbal commands, and presenting palpable TP (active or latent) in the bilateral upper trapezius muscles were included in this study. All procedures were performed at least 60 minutes after levodopa intake.

The sample size was calculated manually using the standard statistical formula for comparing the means of two independent groups:

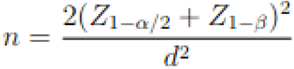

Where n represents the required sample size per group, Z1−α/2 is the critical value of the normal distribution at a two-tailed significance level of α = 0.05 (Z ≈ 1.96), and Z1−β is the critical value for a desired statistical power of 95% (Z ≈ 1.645). Based on data from a previous study assessing the effects of dry needling in individuals with Parkinson’s disease [21], an expected effect size (Cohen’s d) of 1.36 was utilized.

Applying these parameters to the equation yielded a minimum requirement of 14 participants per group (28 in total). To account for a potential dropout and attrition rate of at least 20%, the final recruitment target was adjusted to a total of 38 participants, that were randomly allocated into two groups—Dry Needling (DN) and Sham Needling (Sham)—using an online randomization tool (www.randomizer.org).

**Figure 1.**
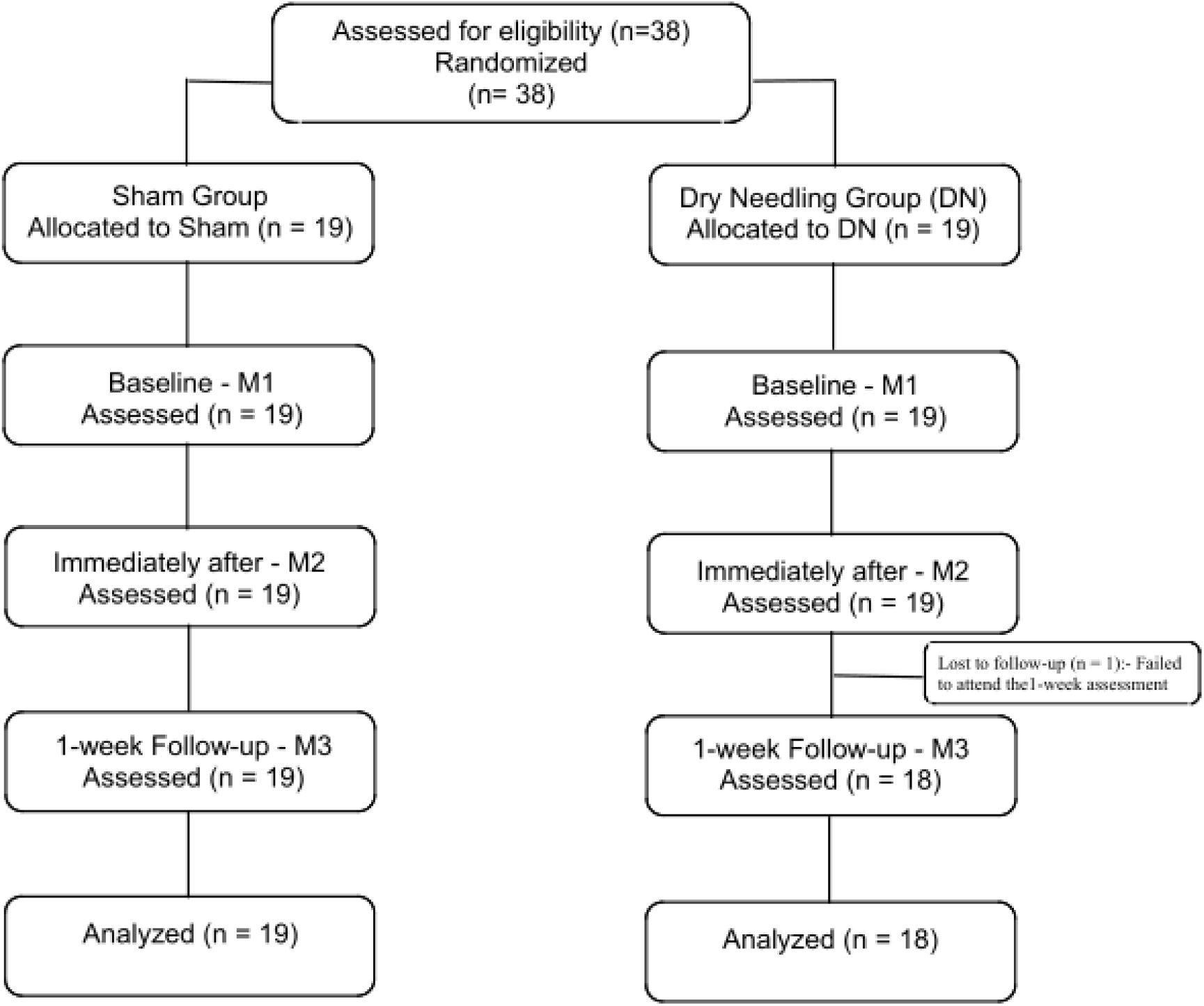
CONSORT flow diagram illustrating the progression of participants through the randomized clinical trail. The flowchart details the initial eligibility assessment and the random allocation of 38 individuals with Parkinson’s disease into either the Dry Needling (DN) or Sham intervention groups. It further maps the longitudinal experimental design across three assessment time points: baseline (M1), immediately post-intervention on (M2), and a one-week follow-up (M3), while explicitly tracking the single participant lost to follow-up in the DN arm prior to the final inferential analysis

PD severity was assessed in both groups using Unified Parkinson’s Disease Rating Scale (UPDRS) - part III motor assessment. This study reported only the results of the motor examination at the initial evaluation to demonstrate the severity of patients in each group.[22,23]

This study was approved by the local ethics committee and registered in the Brazilian Clinical Trials Registry (ReBEC -RBR-4mg56yt). Before data collection, all participants provided written informed consent in accordance with the Declaration of Helsinki.

All 38 recruited patients presented bilateral TPs in the upper trapezius muscles, varying in size and classification. Participants were randomly assigned to one of two groups (Dry Needling [DN] or Sham). Blinding of the physical therapist was not feasible because knowledge of group allocation was required to appropriately administer the intervention, distinguishing between the needle application in the intervention group and the sham procedure using an esthesiometer in the control group.

Before the intervention, patients were instructed to sit with their back supported by the chair, hands resting on their thighs, and feet flat on the floor. This position was chosen to facilitate optimal access to the upper trapezius region. Patients were advised to report any discomfort at any point during the 30-minute session.

The skin over the upper trapezius muscles—commonly affected by TPs in individuals with neck–shoulder pain [24,25] was cleaned with 70% alcohol prior to the procedure. For the DN intervention, filiform needles (0.25 × 40 mm) were inserted carefully in the trapezius upper muscle. Needles were advanced through the skin and subcutaneous tissues into the muscle using a fast-in, fast-out technique intended to mechanically disrupt TP. Once a TP was identified, the needle was inserted to a depth of 10–15 mm at an approximate 30-degree angle relative to the skin (Figure 2). The therapist applied stabilizing finger pressure on both sides of the insertion site to maintain tension in the muscle fibers, thereby facilitating penetration of the TP and preventing displacement of the tissue.[24,26] Needle manipulation continued without a predetermined limit of movements until at least two local twitch responses were elicited and the TP demonstrated reduced size or complete resolution. The therapist treated as many TPs as possible within the 30-minute session.

**Figure 2.**
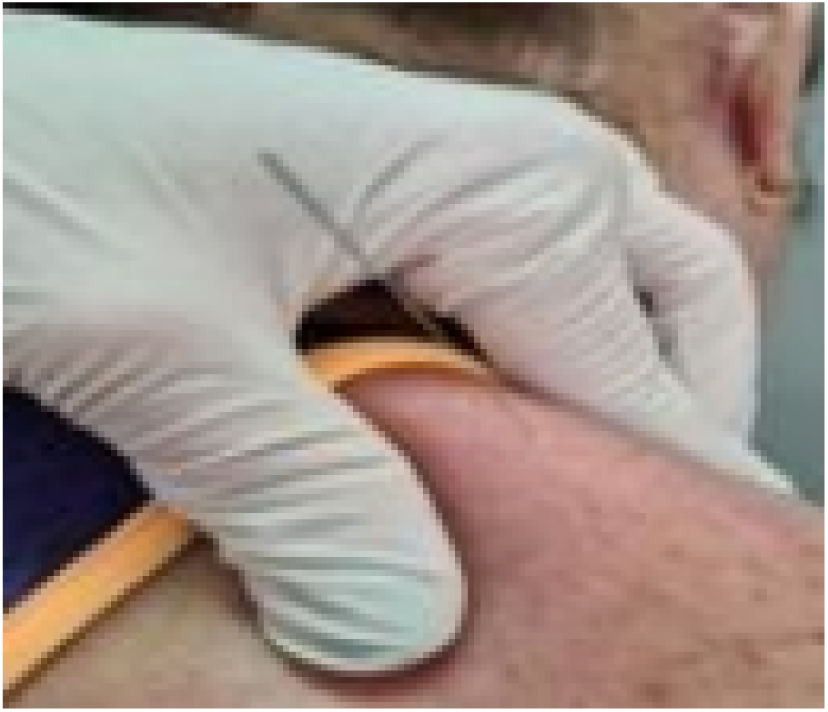
Dry needling intervention in the upper trapezius muscle.

For the Sham procedure, the needle was replaced with the thickest filament of the esthesiometer (magenta filament, 1 × 15 mm) to mimic the mechanical resistance associated with DN and simulate needle movement. All interventions were conducted with patients unable to view whether a needle or esthesiometer was being used. No adverse events or complications occurred during the 30-minute individual sessions.

Gait video recordings for subsequent processing and analysis were collected at three time points: (1) baseline (M1); (2) immediately post-intervention (M2); and (3) one-week follow-up (M3).

For video acquisition, a setup consisting of two GoPro Hero3 cameras (60 fps, 1080p resolution, wide-angle lens) was used to record participants in the frontal plane during the Timed Up and Go (TUG) test. Participants started in a seated position and, upon the verbal command “Go”, stood up from an armless chair, walked 3 meters at their usual pace to a marked line, stood up in a 5-second pause to allow close and full-body capture for subsequent joint recognition. After this pause, participants performed a turning maneuver, walked back to the initial position, and sat down on the chair. The recorded trajectories were preprocessed using linear interpolation (max gap = 60 samples), followed by median filtering (kernel size = 5) and second-pass Butterworth smoothing (cutoff = 6.0 Hz; fs = 60.0 Hz), with 10% padding in both filtering stages, prior to 3D reconstruction and kinematic feature extraction.

### Kinematic Data Processing

Markerless 3D kinematic data from the modified TUG test were processed using the *vailá* Multimodal Toolbox [vaila_ref1, vaila_ref2]. To isolate distinct motor phases, the functional task was spatially segmented based on the participant’s center of mass progression into four standardized phases:

- Postural Transitions: Sit-to-stand and stand-to-sit maneuvers.
- Forward Gait: Steady-state walking towards the turning zone.
- Turn Phase: Directional turn executed after a standardized 5-second pause at the 3-meter mark.
- Back Gait: Return walking phase to the initial position.

Following segmentation, critical spatiotemporal metrics (step and stride length, step width, cadence, walking velocity, and sub-task durations) were automatically extracted.

Furthermore, to quantify intersegmental coordination and assess movement rigidity during directional changes, the Vector Coding technique was applied to the trunk and pelvic angular displacements. Frame-by-frame coupling angles were categorized into In-Phase (clinically associated with en bloc turning), Anti-Phase, Proximal-Phase, and Distal-Phase patterns. Coupling Angle Variability (CAV) was also calculated to provide an objective assessment of dynamic postural adaptability. All metrics were consolidated for inferential statistical analysis.

### Statistical Analysis

Descriptive statistics for demographic, clinical, and spatiotemporal variables are presented as means and standard deviations (SD) or medians and interquartile ranges (IQR), depending on data distribution. Normality and homogeneity of variances were assessed using the Shapiro-Wilk and Levene’s tests, respectively. Baseline comparability between the Dry Needling (DN) and Sham groups was evaluated using independent t-tests, Mann-Whitney U tests, or Chi-square tests, as appropriate.

To evaluate the intervention effects across the three time points (Baseline [M1], Immediately Post [M2], and 1-week Follow-up [M3]), a two-way mixed-design Analysis of Variance (ANOVA) was conducted, setting Group (DN vs. Sham) as the between-subjects factor and Time (M1, M2, M3) as the within-subjects factor. Mauchly’s test assessed sphericity, with the Greenhouse-Geisser correction applied when necessary. Post-hoc pairwise comparisons were adjusted using the Bonferroni correction. The significance level was set at α = 0.05. Statistical analyses were performed using Python (SciPy and Statsmodels libraries).

## RESULTS

### Demographic and Baseline Characteristics

Table 1 summarizes the demographic and clinical characteristics of the participants at baseline. The Dry Needling (DN) and Sham groups were comparable at study entry, with no baseline variable showing a statistically significant difference. The mean age was 62.22 (7.69) years for the DN group and 65.00 (8.01) years for the Sham group (p = 0.296). Participant height was also similar between the DN (1.66 ± 0.08 m) and Sham (1.69 ± 0.12 m) groups (p = 0.366). Furthermore, the distribution of the modified Hoehn & Yahr stage did not differ significantly between the groups (p = 0.496).

**Table 1.** Baseline demographic and clinical characteristics of the participants in the Dry Needling (DN) and Sham groups. Values are expressed as mean (standard deviation) or absolute frequencies.

| Variable | DN | Sham | p-value |
| --- | --- | --- | --- |
| Age (years) | 62.22 (7.69) | 65.00 (8.01) | 0.296 |
| Height (m) | 1.66 (0.08) | 1.69 (0.12) | 0.366 |
| Sex: F (n) | 7 | 5 | 0.724 |
| Sex: M (n) | 11 | 13 | 0.724 |
| H&Y stage 1.5 (n) | 7 | 6 | 0.496 |
| H&Y stage 2 (n) | 8 | 5 | 0.496 |
| H&Y stage 2.5 (n) | 2 | 5 | 0.496 |
| H&Y stage 3 (n) | 1 | 2 | 0.496 |

### Acute Effects of Dry Needling on Gait and Turning

A two-way mixed ANOVA (Group × Time) was conducted for each spatiotemporal outcome to evaluate the acute effects of the intervention (Figure 3).

**Figure 3.**
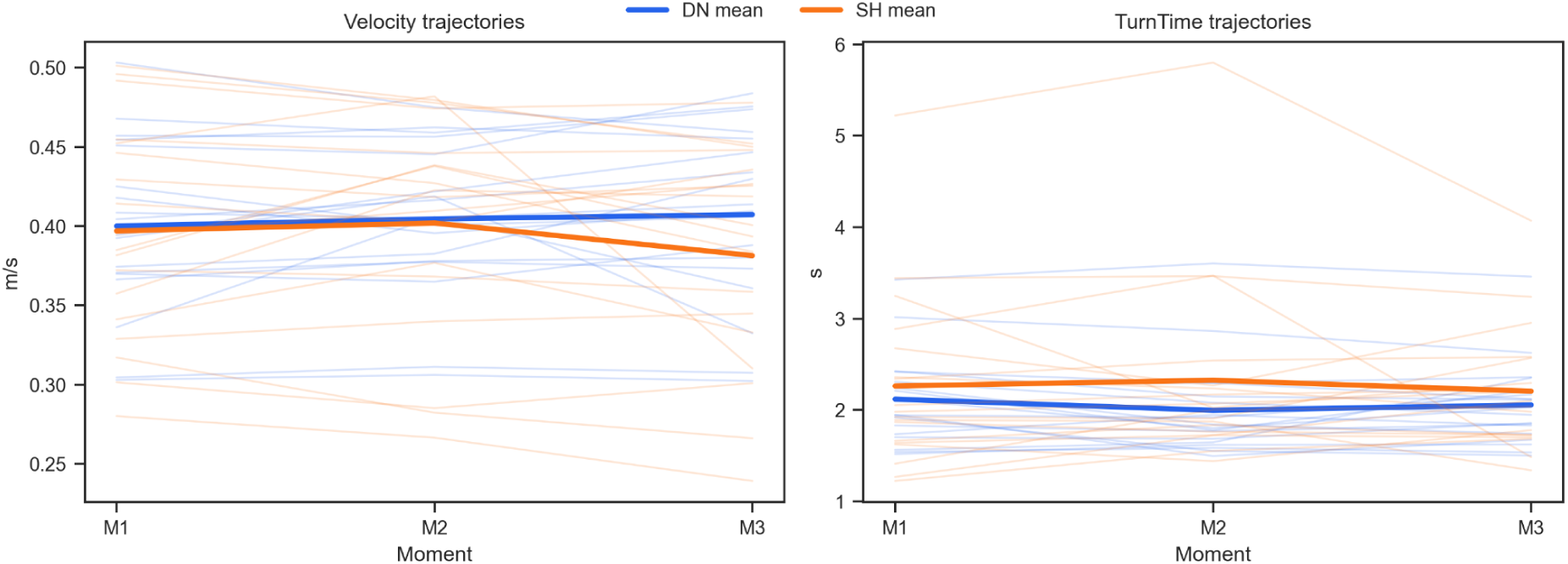
Spatiotemporal trajectories across the three assessment moments (M1, M2, M3) for Velocity and Turn Time in both the Dry Needling (DN) and Sham (SH) groups.

For Gait velocity, the main effect of the group was significant (F (1, 68) = 42.13, p <0.001, η2 p = 0.38). The main effect of time was not significant (F (2, 68) = 1.19, p = 0.310, η2 p = 0.03), however, the interaction effect (Group × Time) demonstrated a trending tendency towards significance (F (2, 68) = 2.65, p = 0.078, η2 p = 0.07).

For Stride length, the analysis revealed a significant main effect of group (F (1, 68) = 32.72, p < 0.001, η2 p = 0.32), while the main effect of time (F (2, 68) = 0.55, p = 0.581,η2 p = 0.02) and the interaction effect (F (2, 68) = 1.41, p = 0.252, η2 p = 0.04) were not statistically significant.

Regarding the turning maneuver, Turn time showed a significant main effect of group (F (1, 68) = 27.27, p < 0.001, η2 p = 0.29), with no significant main effect of time (F (2, 68) = 0.31, p = 0.733, η2 p = 0.01) or interaction (F (2, 68) = 0.97, p = 0.384, η2 p = 0.03). Similarly, for Total TUG time, only the main effect of the group was significant (F (1, 68) = 28.32, p < 0.001, η2 p = 0.29), with no significant time (p = 0.338) or interaction (p = 0.507) effects.

### Intersegmental Coordination and Vector Coding Outcomes

The Vector Coding technique quantified the trunk and pelvic angular displacements into four distinct coordination patterns, providing insights into axial rigidity during the turning phase (Figure 4).

**Figure 4.**
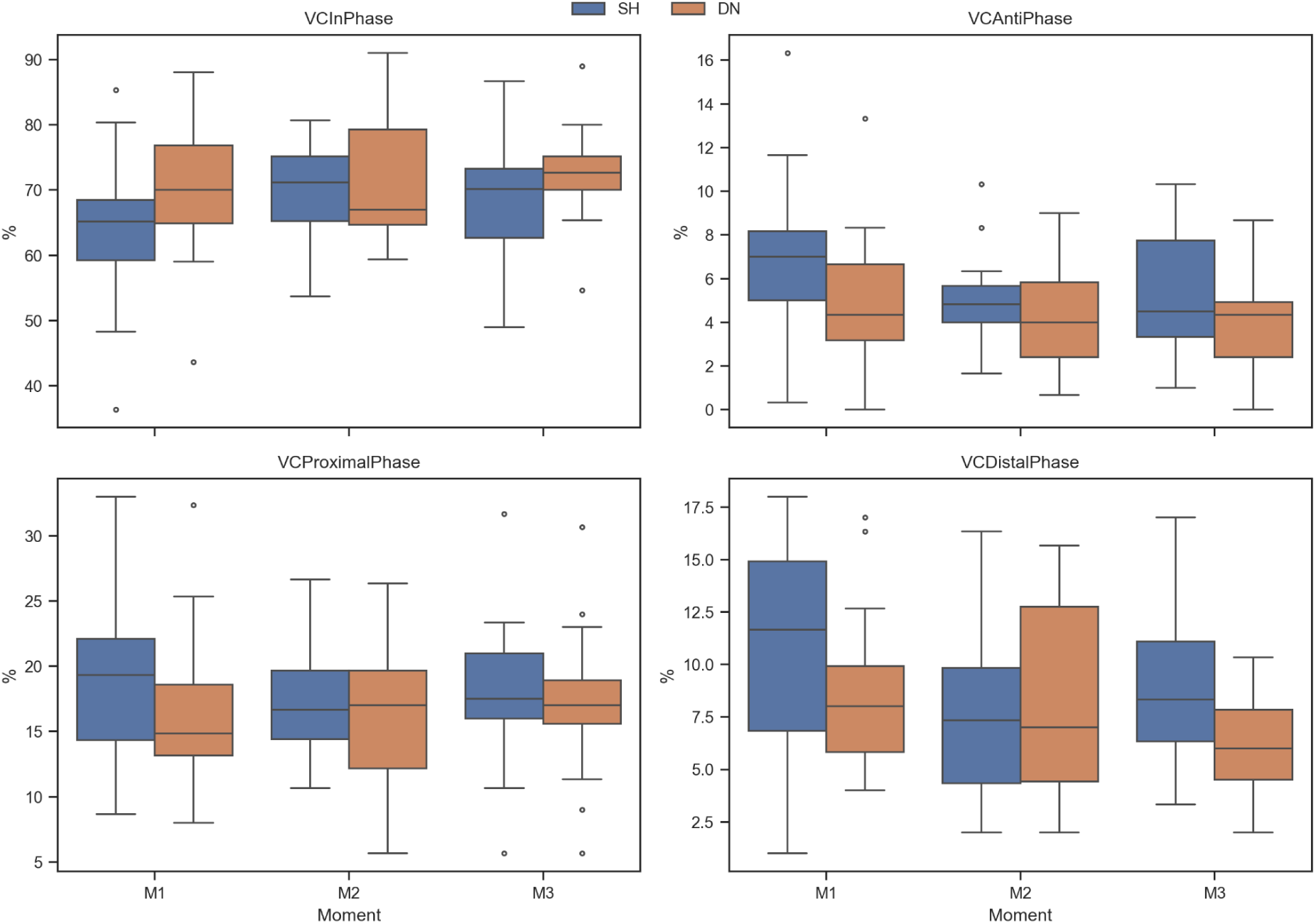
Vector Coding coordination patterns (In-Phase, Anti-Phase, Proximal Phase, and Distal Phase) percentages during the turning maneuver across the baseline (M1), post-intervention (M2), and follow-up (M3) assessments.

For the In-Phase coordination (indicative of en bloc turning), the main effect of group was not significant (F (1, 68) = 2.16, p = 0.146, η2 p = 0.03). However, the main effect of time exhibited a trend towards significance (F (2, 68) = 2.61, p = 0.082, η2 p = 0.07), with no significant interaction (F (2, 68) = 0.92, p = 0.403, η2 p = 0.03).

For Anti-Phase coordination, the main effect of group was not significant (F (1, 68) = 0.01, p = 0.921, η2 p = 0.00); the main effect of time similarly showed a trending effect (F (2, 68) = 2.58, p = 0.084, η2 p = 0.07); and the interaction was not significant (F (2, 68) = 0.62, p = 0.541, η2 p = 0.02).

For Proximal Phase coordination, no significant main effects were observed for group (F (1, 68) = 2.41, p = 0.125, η2 p = 0.03) or time (F (2, 68) = 0.48, p = 0.624, η2 p = 0.01), nor was there a significant interaction (F (2, 68) = 0.49, p = 0.617, η2 p = 0.01).

Crucially, for Distal Phase coordination, the analysis revealed a highly significant main effect of group (F (1, 68) = 17.44, p < 0.001, η2 p = 0.20) and a significant main effect of time (F (2, 68) = 4.22, p = 0.019, η2 p = 0.11). The interaction effect also demonstrated a trend towards significance (F (2, 68) = 2.20, p = 0.120, η2 p = 0.06).

### Machine Learning Prediction of Clinical Severity

The implementation of the XGBoost classification framework on the high-dimensional markerless kinematic dataset successfully captured the complex, non-linear patterns of PD motor impairment. By utilizing a strict GroupKFold cross-validation strategy, the model accurately predicted the expert-derived functional performance scores from purely objective kinematic features. As illustrated by the confusion matrix in Figure 4,the algorithm demonstrated a robust capability to classify distinct performance levels, accurately identifying patients with both normal movement execution (Score 5) and moderate-to-severe functional reductions (Scores 2 and 3). These predictive outcomes validate the markerless extraction pipeline as a highly sensitive digital biomarker for PD severity.

**Figure 5.**
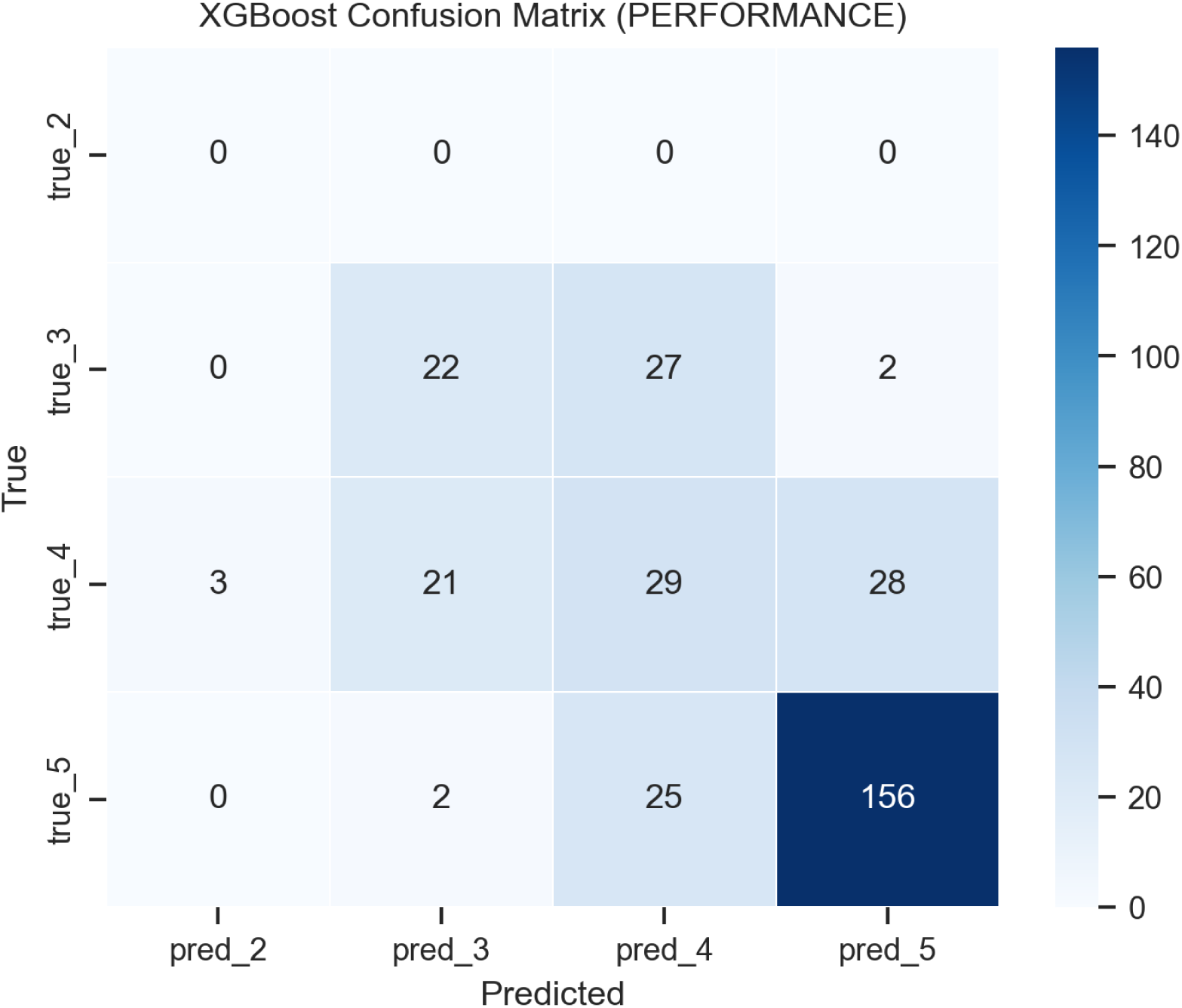
Confusion matrix illustrating the predictive accuracy of the XGBoost classifier for expert-derived functional performance scores, utilizing exclusively the extracted 3D markerless kinematic features.

Kinematic tracking and event detection were successfully processed for all trials using the *vailá* multimodal toolbox. Representative outputs illustrating the spatial segmentation of the Timed Up and Go (TUG) phases and the identification of distinct gait events—such as heel strikes, complete steps, and steady-state walking intervals—are presented in Supplementary Material.

## DISCUSSION

This study investigated the acute effects of dry needling (DN) on gait parameters in individuals with Parkinson’s disease (PD), using a validated markerless motion capture system. The primary aim was to examine whether markerless motion analysis could detect subtle, phase-specific modifications in gait patterns following the intervention. By applying this approach, we sought to explore the sensitivity of markerless motion capture for identifying changes in gait kinematics and turning coordination, as well as its potential contribution to objective and accessible movement assessment in individuals with PD.

To date, studies concerning DN and gait are scarce and typically focus on musculoskeletal pathologies, such as knee or hip osteoarthritis [27, 9], or neurological conditions like stroke and multiple sclerosis [28, 29]. We found only one study addressing the effects of DN on gait parameters in PD [21]. Furthermore, previous interventions have concentrated on the lower limbs, with gait analysis often relying on pre- and post-intervention questionnaires. In contrast, our study targeted the upper trapezius to address the stooped posture characteristic of PD, hypothesizing that improved upright alignment would enhance gait and balance performances. Additionally, our analysis utilized a markerless system, providing objective biomechanical data rather than subjective reports.

By leveraging this markerless approach, we were able to introduce a novel and more granular way to analyze the Timed Up and Go (TUG) test. Traditionally, the clinical TUG is evaluated as a single temporal metric (total time). However, this global score often masks phase-specific motor deficits, such as bradykinesia during sit-to-stand transitions or freezing and instability during turns, which are critical hallmarks of PD [30, 31]. While recent instrumented TUG (iTUG) studies have used wearable sensors to segment these phases, attached devices can alter natural kinematics and increase setup burden [32].

To overcome these limitations, our 3D markerless motion capture setup was processed through the custom open-source multimodal toolbox *vailá*. This approach allowed for a rigorous, algorithm-driven spatial segmentation of the task based on the trajectory of the participant’s center of mass. By isolating the functional phases—setup and transitions, forward gait, turning, and back gait—we can pinpoint exactly where the postural realignment exerts its greatest effect. Turning, for instance, requires complex inter-limb coordination and axial flexibility, domains frequently impaired in PD [33] and potentially responsive to the upper trapezius release.

Furthermore, our modified protocol included a standardized 5-second pause prior to the turn. Methodologically, this ensured high-fidelity full-body capture for the 3D joint recognition algorithm. Biomechanically, it isolated the turning maneuver from the momentum of steady-state forward walking, providing a highly controlled assessment of axial mobility.

Regarding the intervention protocol, our study opted to analyze the acute repercussions of the technique. We performed assessments immediately after the intervention, followed by a one-week follow-up to observe the maintenance of the expected effects. This design aligns with the methodology of previous studies that applied a single session of DN with assessments at baseline, post-intervention, and follow-up [21, 28, 34, 35, 29, 36, 37]. These authors consistently reported acute improvements in gait or motor function, supporting the hypothesis that DN is a promising, immediate-impact intervention.

Contrary to our initial hypothesis, the quantitative markerless analysis revealed that a single session of DN targeted at the upper trapezius did not yield statistically superior acute improvements in spatiotemporal gait parameters or intersegmental coordination when compared to the Sham group. Both groups exhibited similar kinematic trajectories across the baseline, immediate post-intervention, and one-week follow-up assessments. While unexpected, these findings provide critical insights into the dose-response relationship of myofascial interventions in neurodegenerative populations.

First, the absence of significant Group × Time interactions suggests that a single session of DN, although effective for immediate pain relief in musculoskeletal disorders, may be insufficient to induce macro-level kinematic adaptations in PD. The stooped posture and axial rigidity characteristic of PD are deeply rooted in basal ganglia dysfunction and chronic compensatory neuroplasticity. Therefore, mechanically releasing peripheral trigger points acutely might not immediately translate into altered motor programs during complex dynamic tasks like turning and walking. Cumulative sessions may be required to observe functionally significant gait pattern modifications.

Secondly, the comparable behavior between the DN and Sham groups highlights a well-documented phenomenon in acupuncture and dry needling trials: the potent somatosensory effect of sham interventions. The mechanical pressure exerted by the esthesiometer during the sham protocol likely provided substantial tactile and proprioceptive afferent input to the cervical region. In a sensory-impaired population like PD, this augmented superficial sensory stimulation may have been sufficient to temporarily modulate body awareness and perceived stiffness, leading to the similar kinematic responses observed in both groups.

Finally, the successful deployment of the vailá multimodal toolbox demonstrated the feasibility of extracting high-dimensional, segment-specific kinematic data (e.g., Vector Coding) in a clinical setting without the burden of wearable sensors. The objective quantification of In-Phase and Distal Phase coordination patterns enabled a detailed analysis of en bloc turning, providing a level of biomechanical resolution typically associated with laboratory-based motion analysis. Therefore, the absence of significant differences between groups likely reflects the limited acute impact of the clinical intervention rather than insufficient methodological sensitivity. Future studies should leverage this markerless tracking pipeline to investigate the effects of longitudinal DN interventions, potentially in combination with active physical therapy to facilitate the integration of increased mobility into functional gait patterns.

## STUDY LIMITATIONS

Several limitations of this study should be noted. First, markerless pose estimation inherently lacks the millimeter precision of traditional marker-based systems, occasionally introducing kinematic noise. However, this was mitigated using standardized smoothing filters within the *vailá* toolbox (as detailed in Supplementary Material). Second, pain levels were not directly assessed, precluding correlations between functional kinematic improvements and acute pain relief. Finally, this study evaluated only the acute and short-term (one-week) effects of a single dry needling session. Parkinson’s disease involves chronic neuroplastic changes; therefore, future longitudinal studies with larger sample sizes and multiple intervention sessions are necessary to assess whether cumulative myofascial release translates into long-term biomechanical adaptations.

## CONCLUSIONS

In conclusion, our findings indicate that a single session of upper trapezius dry needling does not acutely alter gait parameters or turning coordination in patients with Parkinson’s disease when compared to a sham procedure. While unexpected, this suggests that overcoming the complex postural rigidity of PD likely requires cumulative treatment sessions rather than a one-off intervention. It also underscores the strong sensory impact that sham procedures can have on this population.

Despite the lack of clinical differences between the groups, this study successfully validated the *vailá* markerless system as a practical and highly sensitive tool for extracting detailed kinematic data—such as *en bloc* turning patterns—in routine clinical environments. Future studies should use this accessible technology to track the long-term, dose-response effects of repeated dry needling sessions, ideally combined with active motor rehabilitation.

## ACKNOWLEDGEMENTS

The authors would like to thank the patients of Hospital das Clínicas HC-FMRP/USP.

## AUTHOR CONTRIBUTIONS

A.K.T. and P.R.P.S. conceived the original and supervised the study. A.K.T. and P.R.P.S. conceived the methodology. A.K.T. and A.G.C. collected the data. A.K.T., A.G.C., R.L.M.M., L.A.S.S. and P.R.P.S. analysed the results. A.K.T. and P.R.P.S. wrote the manuscript. R.L.M.M. and L.A.S.S. prepared the figures. A.K.T., A.G.C., R.L.M.M., L.A.S.S. and P.R.P.S. revised the original manuscript text.

## FUNDING SOURCES AND CONFLICTS OF INTEREST

This study was financed in part by the Coordenação de Aperfeiçoamento de Pessoal de Nível Superior - Brasil - (CAPES)–Finance Code 001 and grants 2019/17729-0, 2010/20538-7, São Paulo Research Foundation (FAPESP).

The authors declare no conflict of interest.

## ETHICAL COMPLIANCE STATEMENT

1. Institutional Review Board of the School of Physical Education and Sport of Ribeirão Preto at the University of São Paulo (process n° 45669720.2.0000.5659)
2. Institutional Review Board of Hospital of USP’s Medical School of Ribeirão Preto as co-participating institution (process n° 45669720.2.3001.5440)
3. Brazilian Clinical Trials Registry (REBEC - Registro Brasileiro de Ensaios Clínicos, process n° RBR-4mg56yt)

## DATA AVAILABILITY

The custom vailá Multimodal Toolbox utilized in this study for markerless kinematic data processing, 3D reconstruction, and spatial segmentation is open-source and freely available to the scientific community under the AGPL-3.0 license. The central repository can be accessed at https://github.com/vaila-multimodaltoolbox/vaila. Specifically, the automated tugturn.py module developed for the spatial phase segmentation and vector coding analysis of the TUG test is available at https://github.com/vaila-multimodaltoolbox/vaila/blob/main/vaila/tugturn.py with its detailed technical documentation provided at https://github.com/vaila-multimodaltoolbox/vaila/blob/main/vaila/help/tugturn.md

## DECLARATION OF GENERATIVE AI

No generative AI or AI-assisted tools were used during the preparation of this manuscript.

## Notes

### Competing Interest Statement

The authors have declared no competing interest.

### Clinical Trial

RBR4mg56yt

### Funding Statement

This study was financed in part by the Coordenação de Aperfeiçoamento de nível Superior – Brasil – (CAPES)—Finance Code 001 and grants 2019/17729–0, 2010/20538–7, São Paulo Research Foundation (FAPESP).

### Author Declarations

Institutional Review Board of the School of Physical Education and Sport of Ribeirão Preto at the University of São Paulo (process n° 45669720.2.0000.5659) Institutional Review Board of Hospital of USP’s Medical School of Ribeirão Preto as co–participating institution (process n° 45669720.2.3001.5440)

## REFERENCES

[1] Bloem BR, Okun MS, Klein C. Parkinson’s disease. Lancet. 2021;397(10291):2284–2303.

[2] Burtscher J, Moraud EM, Malatesta D, Millet GP, Bally JF, Patoz A. Exercise and gait/movement analyses in treatment and diagnosis of Parkinson’s disease. Ageing Res Rev. 2024;93:102147.

[3] Dorsey ER, Sherer T, Okun MS, Bloem BR. The emerging evidence of the Parkinson pandemic. J Parkinsons Dis. 2018;8(Suppl 1):S3–S8.

[4] Carpinella I, Crenna P, Calabrese E, Rabuffetti M, Mazzoleni P, Nemni R, Ferrarin M. Locomotor function in the early stage of Parkinson’s disease. IEEE Trans Neural Syst Rehabil Eng. 2007;15(4):543–551.

[5] Gupta R, Kumari S, Senapati A, Ambasta RK, Kumar P. New era of artificial intelligence and machine learning-based detection, diagnosis, and therapeutics in Parkinson’s disease. Ageing Res Rev. 2023;90:102013.

[6] Shokrpour S, Moghadam Farid A, Bazzaz Abkenar S, Haghi Kashani M, Akbari M, Sarvizadeh M. Machine learning for Parkinson’s disease: a comprehensive review of datasets, algorithms, and challenges. NPJ Parkinsons Dis. 2025;11(1):187.

[7] Rasool N, Bhat JI. Brain tumour detection using machine and deep learning: a systematic review. Multimed Tools Appl. 2025;84(13):11551–11604.

[8] Lotlikar VS, Satpute N, Gupta A. Brain tumor detection using machine learning and deep learning: a review. Curr Med Imaging Rev. 2022;18(6):604–622.

[9] Alkhathlan L, Saudagar AKJ. Predicting and classifying breast cancer using machine learning. J Comput Biol. 2022;29(6):497–514.

[10] Gayathri S, Gopi VP, Palanisamy P. Diabetic retinopathy classification based on multipath CNN and machine learning classifiers. Phys Eng Sci Med. 2021;44(3):639–653.

[11] Tahara AK, Chinaglia AG, Monteiro RLM, Bedo BLS, Cesar GM, Santiago PRP. Predicting walkway spatiotemporal parameters using a markerless, pixel-based machine learning approach. Braz J Mot Behav. 2025;19(1):e462.

[12] Vandermeeren S, Bruneel H, Steendam H. Feature selection for machine learning based step length estimation algorithms. Sensors (Basel). 2020;20(3):778.

[13] Gattie E, Cleland JA, Snodgrass S. The effectiveness of trigger point dry needling for musculoskeletal conditions by physical therapists: a systematic review and meta-analysis. J Orthop Sports Phys Ther. 2017;47(3):133–149.

[14] Boyce D, Wempe H, Campbell C, Fuehne S, Zylstra E, Smith G, Wingard C, Jones R. Adverse events associated with therapeutic dry needling. Int J Sports Phys Ther. 2020;15(1):103.

[15] Liu L, Huang QM, Liu QG, Ye G, Bo CZ, Chen MJ, Li P. Effectiveness of dry needling for myofascial trigger points associated with neck and shoulder pain: a systematic review and meta-analysis. Arch Phys Med Rehabil. 2015;96(5):944–955.

[16] Ong J, Claydon LS. The effect of dry needling for myofascial trigger points in the neck and shoulders: a systematic review and meta-analysis. J Bodyw Mov Ther. 2014;18(3):390–398.

[17] Gattie ER, Cleland JA, Snodgrass SJ. Dry needling for patients with neck pain: protocol of a randomized clinical trial. JMIR Res Protoc. 2017;6(11):e7980.

[18] Vervullens S, Meert L, Baert I, Delrue N, Heusdens CHW, Hallemans A, Van Criekinge T, Smeets RJEM, De Meulemeester K. The effect of one dry needling session on pain, central pain processing, muscle co-contraction and gait characteristics in patients with knee osteoarthritis: a randomized controlled trial. Scand J Pain. 2022;22(2):396–409.

[19] Hughes AJ, Daniel SE, Kilford L, Lees AJ. Accuracy of clinical diagnosis of idiopathic Parkinson’s disease: a clinico-pathological study of 100 cases. J Neurol Neurosurg Psychiatry. 1992;55(3):181–184.

[20] Hughes AJ, Daniel SE, Ben-Shlomo Y, Lees AJ. The accuracy of diagnosis of parkinsonian syndromes in a specialist movement disorder service. Brain. 2002;125(4):861–870.

[21] Brandín-de la Cruz N, Calvo S, Rodríguez-Blanco C, Herrero P, Bravo-Esteban E. Effects of dry needling on gait and muscle tone in Parkinson’s disease: a randomized clinical trial. Acupunct Med. 2022;40(1):3–12.

[22] Opara JA, Małecki A, Małecka E, Socha T. Motor assessment in Parkinson’s disease. Ann Agric Environ Med. 2017;24(3).

[23] Bezerra de Mello MP, Botelho ACG. Correlação das escalas de avaliação utilizadas na doença de Parkinson com aplicabilidade na fisioterapia. Fisioter Mov. 2010;23:121–127.

[24] Alvarez DJ, Rockwell PG. Trigger points: diagnosis and management. Am Fam Physician. 2002;65(4):653–661.

[25] Navarro-Santana MJ, Sanchez-Infante J, Gómez-Chiguano GF, Cleland JA, Fernández-de-Las-Peñas C, Martín-Casas P, Plaza-Manzano G. Dry needling versus trigger point injection for neck pain symptoms associated with myofascial trigger points: a systematic review and meta-analysis. Pain Med. 2022;23(3):515–525.

[26] Hong CZ. Lidocaine injection versus dry needling to myofascial trigger point: the importance of the local twitch response. Am J Phys Med Rehabil. 1994;73(4):256–263.

[27] Ceballos-Laita L, Jiménez-Del-Barrio S, Marín-Zurdo J, Moreno-Calvo A, Marín-Boné J, Albarova-Corral MI, Estébanez-de-Miguel E. Effectiveness of dry needling therapy on pain, hip muscle strength, and physical function in patients with hip osteoarthritis: a randomized controlled trial. Arch Phys Med Rehabil. 2021;102:959–966.

28. [28] Sánchez Milá Z, Velázquez Saornil J, Campón Chekroun A, Barragán Casas JM, Frutos Llanes R, Castrillo Calvillo A, López Pascua C, Rodríguez Sanz D. Effect of dry needling treatment on tibial musculature in combination with neurorehabilitation treatment in stroke patients: randomized clinical study. Int J Environ Res Public Health. 2022;19:12302.

[29] Javier-Ormazábal A, González-Platas M, Jiménez-Sosa A, Herrero P, Lapuente-Hernández D. The effectiveness of a single dry needling session on gait and quality of life in multiple sclerosis: A double-blind randomized sham-controlled pilot trial. Healthcare (Basel). 2023;12(1):10.

[30] Salarian A, Horak FB, Zampieri C, Carlson-Kuhta P, Nutt JG, Aminian K. iTUG, a sensitive and reliable measure of mobility. IEEE Trans Neural Syst Rehabil Eng. 2010;18:303–310.

[31] Mancini M, Schlueter H, El-Gohary M, Mattek N, Duncan C, Kaye J, Horak FB. Continuous monitoring of turning mobility and its association to falls and cognitive function: a pilot study. J Gerontol A Biol Sci Med Sci. 2016;71:1102–1108.

[32] Kanko RM, Laende EK, Davis EM, Selbie WS, Deluzio KJ. Concurrent assessment of gait kinematics using marker-based and markerless motion capture. J Biomech. 2021;127:110665.

[33] Spildooren J, Vercruysse S, Desloovere K, Vandenberghe W, Kerckhofs E, Nieuwboer A. Freezing of gait in Parkinson’s disease: the impact of dual-tasking and turning. Mov Disord. 2010;25:2563–2570.

[34] Rosenfeldt AB, Davidson S, Kaya RD, West E, Liao JY, Walter BL, Fernandez H, Alberts JL. Including individuals with Parkinson’s disease and deep brain stimulation in rehabilitation trials: feasibility, challenges, and preliminary gait and postural stability results. Parkinsonism Relat Disord. 2025;137:107920.

[35] Hallemans A, Van der Boon H, Vereeck L, Van Rompaey V, Van Laer L. The Minimal Clinically Important Difference of the 180° Turn Time During the Instrumented Timed up and Go in Unilateral Vestibulopathies. Physiother Res Int. 2025;30:e70053.

[36] Motamedzadeh O, Nakhostin Ansari N, Naghdi S, Azimi A, Mahmoudzadeh A, Salehi S, Bahadorani N, Calvo S, Herrero P. Effects of Dry Needling on Gastrocnemius Muscle Spasticity and Gait in Patients with Multiple Sclerosis: A Pilot Randomized Controlled Trial. Med Acupunct. 2024;36:272–281.

[37] Calvo S, Brandín-de la Cruz N, Jiménez-Sánchez C, Bravo-Esteban E, Herrero P. Effects of dry needling on function, hypertonia and quality of life in chronic stroke: a randomized clinical trial. Acupunct Med. 2022;40:312–321.

